# Changes in paediatric respiratory infections at a UK teaching hospital 2016-2021; impact of the SARS-CoV-2 pandemic

**DOI:** 10.1101/2021.10.13.21264956

**Authors:** Sheila F. Lumley, Nicholas Richens, Emily Lees, Jack Cregan, Elizabeth Kalimeris, Sarah Oakley, Marcus Morgan, Shelley Segal, Moya Dawson, A. Sarah Walker, David W. Eyre, Derrick W. Crook, Sally Beer, Alex Novak, Nicole E. Stoesser, Philippa C. Matthews

**Affiliations:** Oxford University Hospitals NHS Foundation Trust, Oxford, UK; Nuffield Department of Medicine, University of Oxford, Oxford, UK; Department of Paediatrics, University of Oxford, Oxford UK; NIHR Oxford Biomedical Research Centre, University of Oxford, Oxford, UK; NIHR Health Protection Research Unit in Healthcare Associated Infections and Antimicrobial Resistance, University of Oxford, Oxford, UK; Nuffield Department of Population Health, University of Oxford, Oxford, UK; Big Data Institute, University of Oxford, Oxford, UK

**Keywords:** respiratory virus, respiratory tract infection, paediatric, SARS-CoV-2, respiratory syncytial virus, influenza, rhinovirus, coinfection

## Abstract

**Objective:** To describe the impact of the SARS-CoV-2 pandemic on the incidence of paediatric viral respiratory tract infection in Oxfordshire, UK.

**Methods:** Data on paediatric Emergency Department (ED) attendances (0-15 years inclusive), respiratory virus testing, vital signs and mortality at Oxford University Hospitals were summarised using descriptive statistics.

**Results:** Between 1-March-2016 and 30-July-2021, 155,056 ED attendances occurred and 7,195 respiratory virus PCRs were performed. Detection of all pathogens was suppressed during the first national lockdown. Rhinovirus and adenovirus rates increased when schools reopened September-December 2020, then fell, before rising in March-May 2021. The usual winter RSV peak did not occur in 2020/21, with an inter-seasonal rise (32/1,000 attendances in 0-3yr olds) in July 2021. Influenza remained suppressed throughout. A higher Paediatric Early Warning Score (PEWS) was seen for attendees with adenovirus during the pandemic compared to pre-pandemic (p=0.04, Mann-Witney U test), no other differences in PEWS were seen.

**Conclusions:** SARS-CoV-2 caused major changes in the incidence of paediatric respiratory viral infection in Oxfordshire, with implications for clinical service demand, testing strategies, timing of palivizumab RSV prophylaxis, and highlighting the need to understand which public health interventions are most effective for preventing respiratory virus infections.

## INTRODUCTION

Respiratory tract infections (RTI) represent a major global disease burden in children, particularly in children under five years of age. Lower RTI are one of the top ten causes of mortality and morbidity considered by the World Health Organisation (WHO)(1). Although many RTIs are mild and self-limiting, they remain one of the commonest reasons for primary care consultation, attendance in emergency departments, hospital admission and antibiotic prescribing. In the UK, incidence of upper RTI is around 300,000 cases per 100,000 people per year(2), with children <5 years experiencing as many as 10 infections/year(3).

Prior to the SARS-CoV-2 pandemic, common causes of RTI included bacterial pathogens (e.g. *Streptococcus pneumoniae, Haemophilus influenzae* [in settings with low vaccination rates]) and viruses such as respiratory syncytial virus (RSV), influenza, parainfluenza, adenoviruses, rhinovirus, respiratory enteroviruses and non-SARS-CoV-2 human coronaviruses (hCov), with viral infections accounting for ∼40-50% of RTI presentations. Symptomatic paediatric SARS-CoV-2 infection remains uncommon, accounting for <2% of COVID-19 cases in England, with few severe infections and deaths (1-5/100,000 paediatric infections requiring admission, and even fewer requiring intensive care admission). Particular attention has been related to rare presentations in children with paediatric multisystem inflammatory syndrome temporally-associated with SARS-CoV-2 (PIMS-TS) which can be life-threatening, with 44% of PIMS-TS admissions in the UK requiring intensive care(4–6).

Notably however, the SARS-CoV-2 pandemic has resulted in major observed changes to the wider epidemiology of RTIs, driven by social distancing, use of face coverings and periods of lockdown including closure of daycare and educational settings. This has led to reduced opportunities for virus transmission, and potential shifts in interactions and competition amongst respiratory pathogens. This includes significant reductions in non-SARS-CoV-2-associated RTIs in adult and paediatric populations(7–13). Atypical rebounds and peaks in rhinovirus and RSV infections have been observed following the loosening of social restrictions and reopening of educational settings, likely partly driven by waning population immunity given the lack of exposure to these pathogens during 2020(14–23). This has been particularly prominent in the case of RSV, with presentations reported in a significantly older population of children than would usually be affected(23).

Here, we describe the impact of the COVID-19 pandemic on the incidence of paediatric viral RTI in Oxfordshire, UK.

## MATERIALS AND METHODS

Oxford University Hospitals (OUH) is a tertiary referral centre consisting of four hospital sites in Oxfordshire: three hospitals in Oxford serving a population of 655,000, and a fourth in Banbury with a catchment population of around 150,000. Emergency Departments (ED) in Oxford and Banbury see children between the age of 0-15 years.

Data on paediatric ED attendances, baseline characteristics such as age, gender, ethnicity, index of multiple deprivation (IMD), hospital admission (general wards vs. critical care), respiratory virus test results (within the first 24 hours after arrival at ED), vital signs (all heart rate, respiratory rate and oxygen saturation readings recorded in the ED department) and mortality (within 14 days of a positive respiratory virus test) were sourced from the Infections in Oxfordshire Research Database (IORD, https://oxfordbrc.nihr.ac.uk/research-themes-overview/antimicrobial-resistance-and-modernising-microbiology/infections-in-oxfordshire-research-database-iord/).

During the study period, respiratory virus testing was performed according to local protocols, detailed in table 1. Platforms used are detailed in supplementary table 1.

**Table 1:** Paediatric respiratory virus testing policies during study period. ^*^ Respiratory virus targets as follows: Flu A/B/RSV PCR - Influenza A, Influenza B, Respiratory syncytial virus PCR Biofire multiplex respiratory PCR - BioFire FilmArray Respiratory Panel test - Influenza A & B, parainfluenza 1–4, RSV, human enterovirus/rhinovirus, coronaviruses 229E, HKU1, NL63, OC43, adenovirus, MERS-CoV, human metapneumovirus *(SARS-CoV-2 included from October 2020 as part of the BioFire FilmArray Respiratory Panel test 2*.*1)*

| Date | Test* | Policy |
| --- | --- | --- |
| March 2016 - Jan 2020 | Flu A/B/RSV PCR | Only test if changes clinical management<br>All suspected febrile neutropenia during Flu/RSV season |
|  | Biofire multiplex respiratory PCR | Admission to ITU/HDU with a respiratory illness<br>Suspected febrile neutropenia - if URTI symptoms<br>OR at clinician's discretion |
| Feb 2020 - June 2021 | SARS-CoV-2 PCR | Child requiring admission for at least one night AND clinical evidence of pneumonia/pneumonitis |
|  | Flu A/B/RSV PCR | Only test if changes clinical management<br>All suspected febrile neutropenia during Flu/RSV season |
|  | Biofire multiplex respiratory PCR | Admission to ITU/HDU with a respiratory illness<br>OR at clinician's discretion |
| July 2021 | Biofire multiplex respiratory PCR | Admission to ITU/HDU with a respiratory illness<br>OR requiring an aerosol generating procedure<br>OR at clinician's discretion |
|  | Flu A/B/RSV & SARS-CoV-2 PCR | All other children requiring admission |

Data on the strictness of lockdown policies restricting people’s behaviour in England were sourced from Oxford COVID-19 Government Response Tracker (OxCGRT)(24). This ‘stringency index’ is calculated from containment and closure policy indicators capturing information on containment and closure policies such as school closures and restrictions in movement, plus an indicator recording public information campaigns.

Data were categorised into pre-pandemic (before 23-Mar-20) and pandemic (23-Mar-20 onwards) periods. 23-Mar-20 was the date that the UK government announced plans for a national lockdown, selected here to reflect the approximate time when widespread behaviour changes occurred in the UK, rather than the date when SARS-CoV-2 was first introduced into the UK. As only heart rate, respiratory rate and oxygen saturations were reliably available, partial Paediatric Early Warning Scores (PEWS) using the Alder Hey PEWS(25), were used as a marker of disease severity, and calculated based on highest heart rate, highest respiratory rate and lowest oxygen saturation recorded, in attendances where all 3 parameters were available.

### Statistical analysis

Data analysis was performed using R version 4.0.2. For comparisons between pre-pandemic and pandemic periods, Pearson’s Chi-squared test was used for categorical data and Mann-Whitney U test for numeric data. Kruskal-Wallis was used for comparisons between the PEWS for different pathogens.

### Ethical approvals

IORD has generic Research Ethics Committee, Health Research Authority and Confidentiality Advisory Group approvals (REC ref 19/SC/0403; ECC5-017(A)/2009).

## RESULTS

### Demographics of patients presenting to Paediatric ED

Between 1-March-2016 and 30-July-2021, 155,056 Paediatric ED attendances were recorded from 81,339 individuals (supplementary figure 1). The median number of attendances per individual was 1 (IQR 1-2, range 1-52). 41.3% were aged 0-3 years, 37.9% aged 4-11 and 20.7% aged 12-15. The majority of attendances were by males (87,010 (56.1%)) and children of white ethnicity (118,314 (76.3%)). The median Index of Multiple Deprivation score (IMD) was 11.7 (IQR 7.1-20.1, range 0.5-80; a lower score indicates less deprived areas, placing the median score in the 2nd least deprived quintile in England). There were small but statistically significant differences in age, ethnicity and IMD score between pre-pandemic and pandemic periods (Table 2).

**Table 2:**
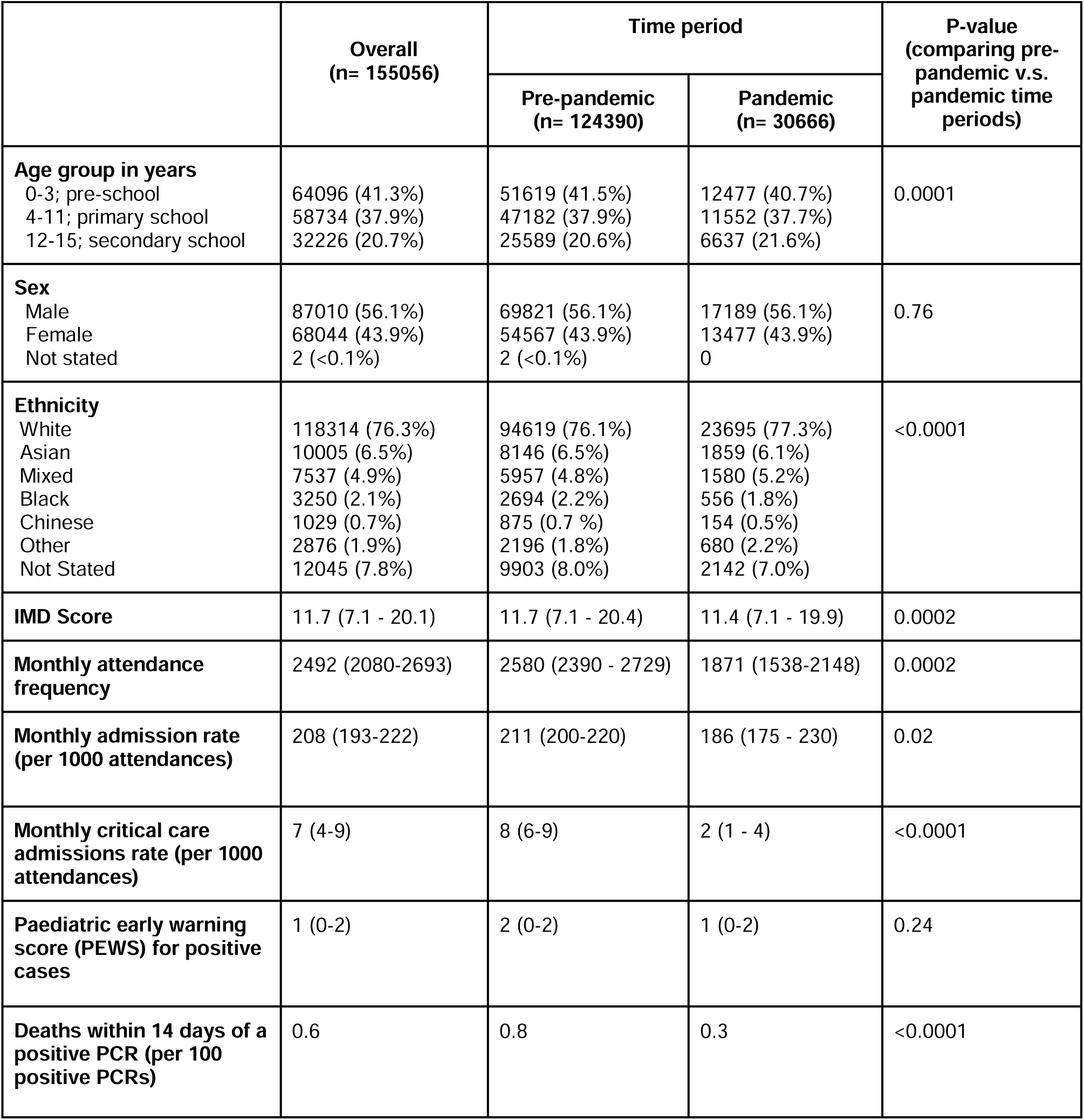
Demographics for 155,056 Paediatric ED attendances between 1-Mar-2016 and 30-July-2021. Median (IQR) or frequency (%) provided. P-values calculated with Pearson’s Chi-squared test for categorical data and Mann-Whitney U test for numeric data. “Pre-pandemic” is defined as prior to 23-Mar-2020, “Pandemic” is defined as 23-Mar-2020 onwards. IMD = index of multiple deprivation PCR = polymerase chain reaction

|  | Overall<br>(n= 155056) | Time period |  | P-value<br>(comparing pre-pandemic v.s. pandemic time periods) |
| --- | --- | --- | --- | --- |
|  |  | Pre-pandemic<br>(n= 124390) | Pandemic<br>(n= 30666) |  |
| **Age group in years**<br>0-3; pre-school<br>4-11; primary school<br>12-15; secondary school | 64096 (41.3%)<br>58734 (37.9%)<br>32226 (20.7%) | 51619 (41.5%)<br>47182 (37.9%)<br>25589 (20.6%) | 12477 (40.7%)<br>11552 (37.7%)<br>6637 (21.6%) | 0.0001 |
| **Sex**<br>Male<br>Female<br>Not stated | 87010 (56.1%)<br>68044 (43.9%)<br>2 (<0.1%) | 69821 (56.1%)<br>54567 (43.9%)<br>2 (<0.1%) | 17189 (56.1%)<br>13477 (43.9%)<br>0 | 0.76 |
| **Ethnicity**<br>White<br>Asian<br>Mixed<br>Black<br>Chinese<br>Other<br>Not Stated | 118314 (76.3%)<br>10005 (6.5%)<br>7537 (4.9%)<br>3250 (2.1%)<br>1029 (0.7%)<br>2876 (1.9%)<br>12045 (7.8%) | 94619 (76.1%)<br>8146 (6.5%)<br>5957 (4.8%)<br>2694 (2.2%)<br>875 (0.7 %)<br>2196 (1.8%)<br>9903 (8.0%) | 23695 (77.3%)<br>1859 (6.1%)<br>1580 (5.2%)<br>556 (1.8%)<br>154 (0.5%)<br>680 (2.2%)<br>2142 (7.0%) | <0.0001 |
| **IMD Score** | 11.7 (7.1 - 20.1) | 11.7 (7.1 - 20.4) | 11.4 (7.1 - 19.9) | 0.0002 |
| **Monthly attendance frequency** | 2492 (2080-2693) | 2580 (2390 - 2729) | 1871 (1538-2148) | 0.0002 |
| **Monthly admission rate (per 1000 attendances)** | 208 (193-222) | 211 (200-220) | 186 (175 - 230) | 0.02 |
| **Monthly critical care admissions rate (per 1000 attendances)** | 7 (4-9) | 8 (6-9) | 2 (1 - 4) | <0.0001 |
| **Paediatric early warning score (PEWS) for positive cases** | 1 (0-2) | 2 (0-2) | 1 (0-2) | 0.24 |
| **Deaths within 14 days of a positive PCR (per 100 positive PCRs)** | 0.6 | 0.8 | 0.3 | <0.0001 |

Pre-pandemic, a median of 2,580 attendances were recorded per month, with peaks in attendance each November in the 0-3 year age group. A median of 211 cases (8.2% of attendances) were admitted per month, 8 (0.3%) to critical care. During the pandemic, median attendance was lower at 1,871 per month (p = 0.0002), and the usual seasonal winter peaks were lost, with an atypical summer peak in attendance in June/July 2021 (>1,250 attendances per month, compared to usual June/July attendance rates of ∼900/month). A median of 186 cases were admitted per month (10% of attendances), 2 (0.1%) to critical care (p=0.02 and p<0.0001 compared to pre-pandemic, respectively) (Table 2, Figure 1A).

**Figure 1:**
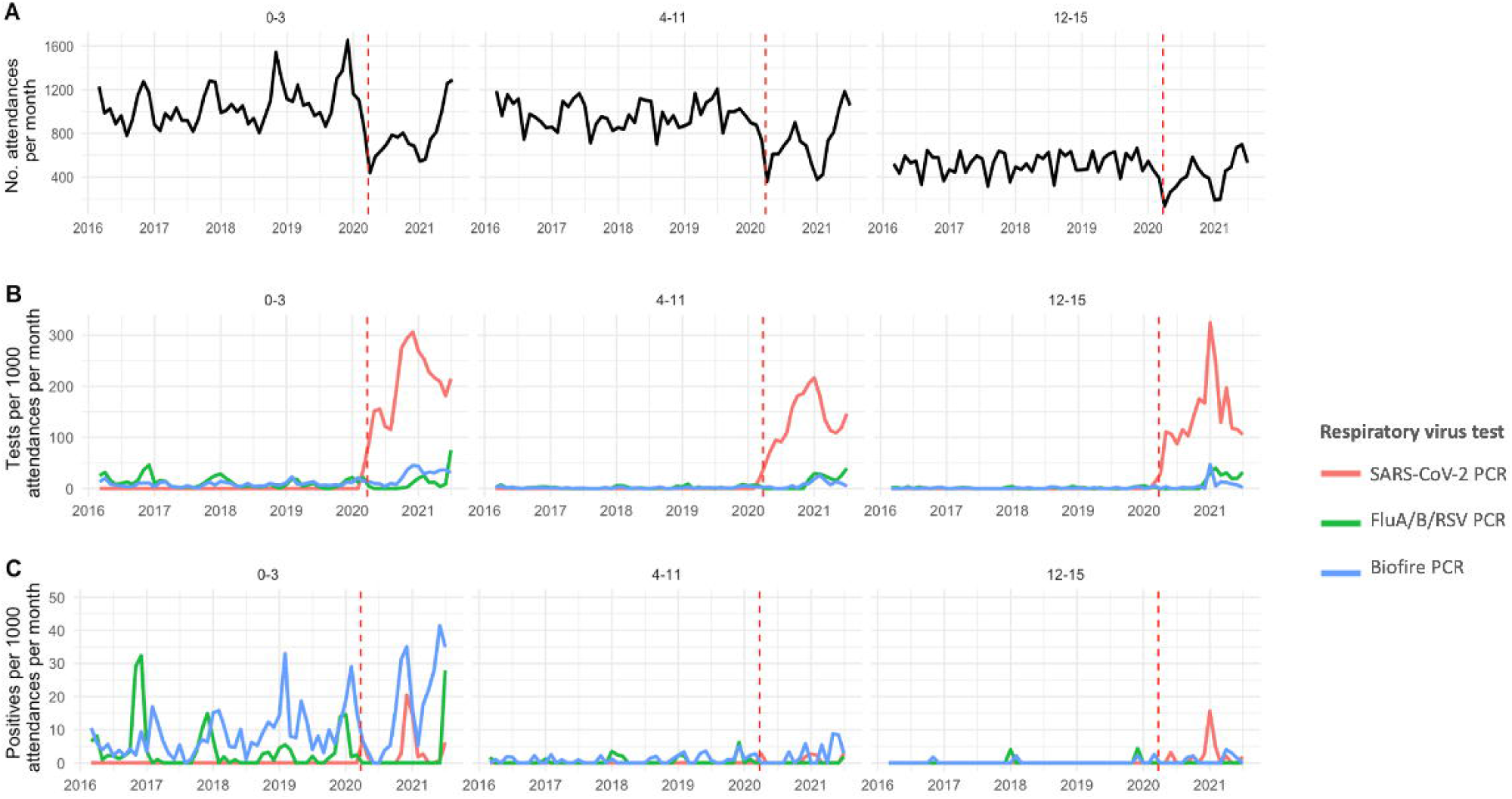
Paediatric attendance, respiratory virus testing and positivity rates over time. A) Rate of paediatric ED attendances, B) Rate of tests per 1000 attendances per month, C) Rate of positives per 1000 attendances per month. Left hand panels show 0-3 years, central panels 4-11 years and right hand panels 12-15 years. Red dashed line indicates the start of the pandemic period, defined here as March 2020. SARS-CoV-2 = SARS-CoV-2 specific PCR, Biofire PCR = BioFire multiplex respiratory PCR.

### Respiratory virus testing patterns over time

A total of 7,195 respiratory virus tests were performed on children during the period observed (1,179 FluA/B/RSV PCR, 1,005 Biofire respiratory multiplex PCR and 5,011 SARS-CoV-2 PCR). Respiratory virus testing rates varied by assay and time (Figure 1B); a median of 4.5 Influenza A/B/RSV PCRs/1,000 attendances were performed pre-pandemic, compared to 7.2/1,000 attendances during the pandemic (p = 0.95), a median of 3.9 Biofire multiplex PCR/1,000 attendances were performed pre-pandemic, compared to 16.7/1,000 attendances during the pandemic (p<0.0001). No SARS-CoV-2 PCRs were performed pre-pandemic, compared to 163/1,000 attendances during the pandemic. Demographic differences between those tested vs. not tested are shown in supplementary table 2.

Prior to the pandemic the majority of tests (1,119/1,355; 83%) were performed on 0-3 year olds, 182/1,355 (13%) on 4-11 year olds and 54/1,355 (4%) on 12-15 year olds. During the pandemic testing increased in the older age groups; 3,039/5,840 (49%) of tests were performed on 0-3 year olds, 1,773/5,840 (30%) on 4-11 year olds and 1,028/5,840 (18%) on 12-15 year olds, reflecting changes in attendance patterns and testing policies over time.

### Respiratory virus detection rates over time

One or more respiratory viruses was identified on 1,128/7,195 (16%) respiratory virus tests, on 935/155,056 ED attendances. The distribution of viruses in pre-pandemic and pandemic periods are shown in Figure 2 and Supplementary table 3. Rates of pathogen detection varied over the study period and by assay performed (Figure 1C). The first case of paediatric SARS-CoV-2 in this study was seen on 24-Mar-20, and in the period since the start of the pandemic it has accounted for 15% of respiratory viruses diagnosed in patients attending Paediatric ED.

**Figure 2:**
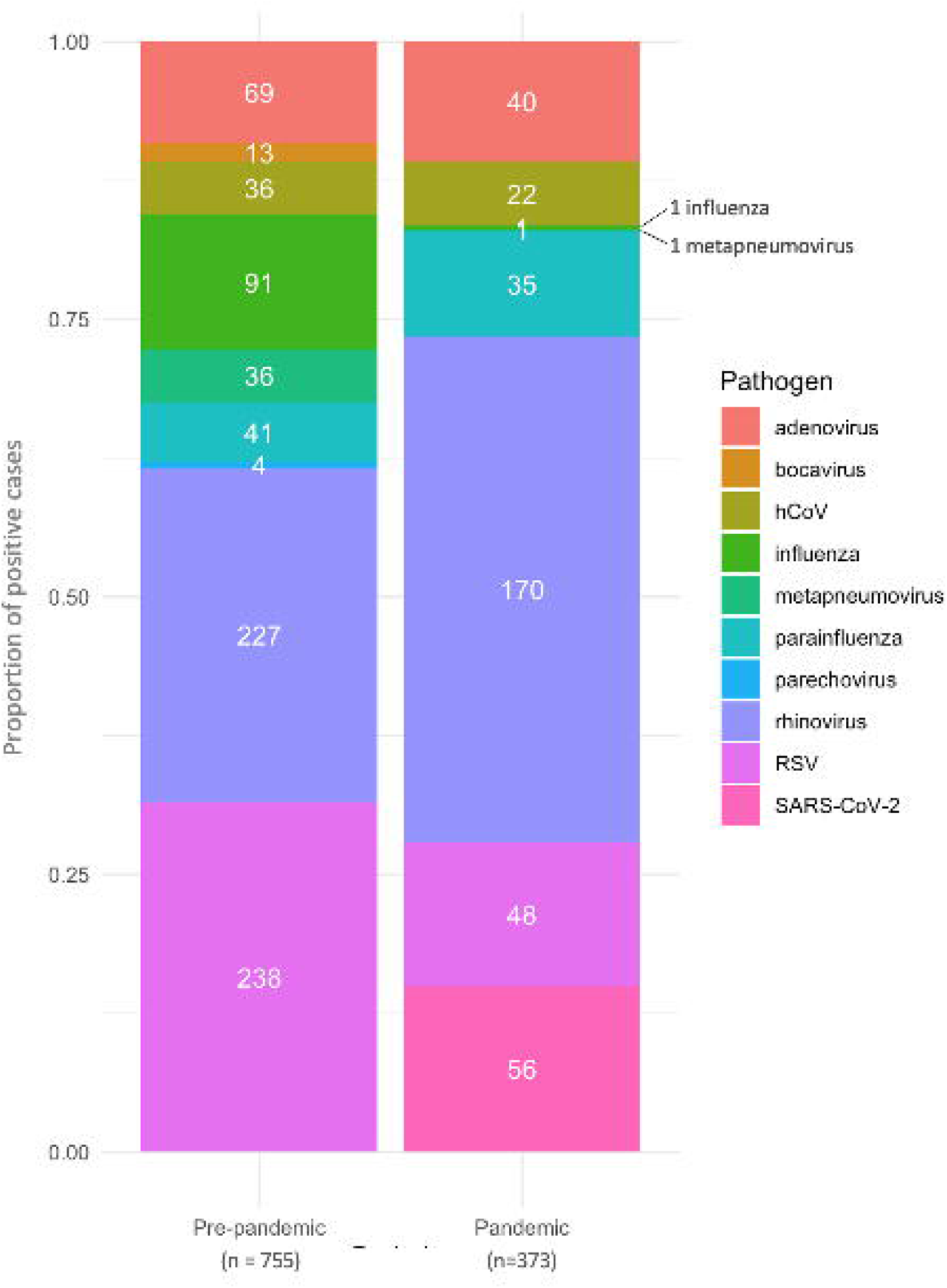
Respiratory virus detection in children age 0-15 pre- and during the SARS-CoV-2 pandemic. Stacked bars represent proportions of pathogens during each period. Frequency of individual pathogens are shown in white text, with totals for each period in the x-axis legend. Respiratory viruses were detected using i) Influenza A/B/RSV PCR, ii) Biofire respiratory multiplex PCR, iii) SARS-CoV-2 PCR or iv) Cepheid Flu A/B/RSV/SARS-CoV-2 (see supplementary table 1).

Pre-pandemic, seasonal peaks of RSV and influenza A/B were observed in September-February and December-March respectively, and to a lesser extent with hCoV in December-March. Rhinovirus and adenovirus exhibited less seasonal variation, with rising case numbers in February in some years.

Numbers of parainfluenza and metapneumovirus were too low to detect seasonal variation. The highest incidence of diagnoses was seen in the 0-3 year olds for all pathogens (Figure 3).

**Figure 3:**
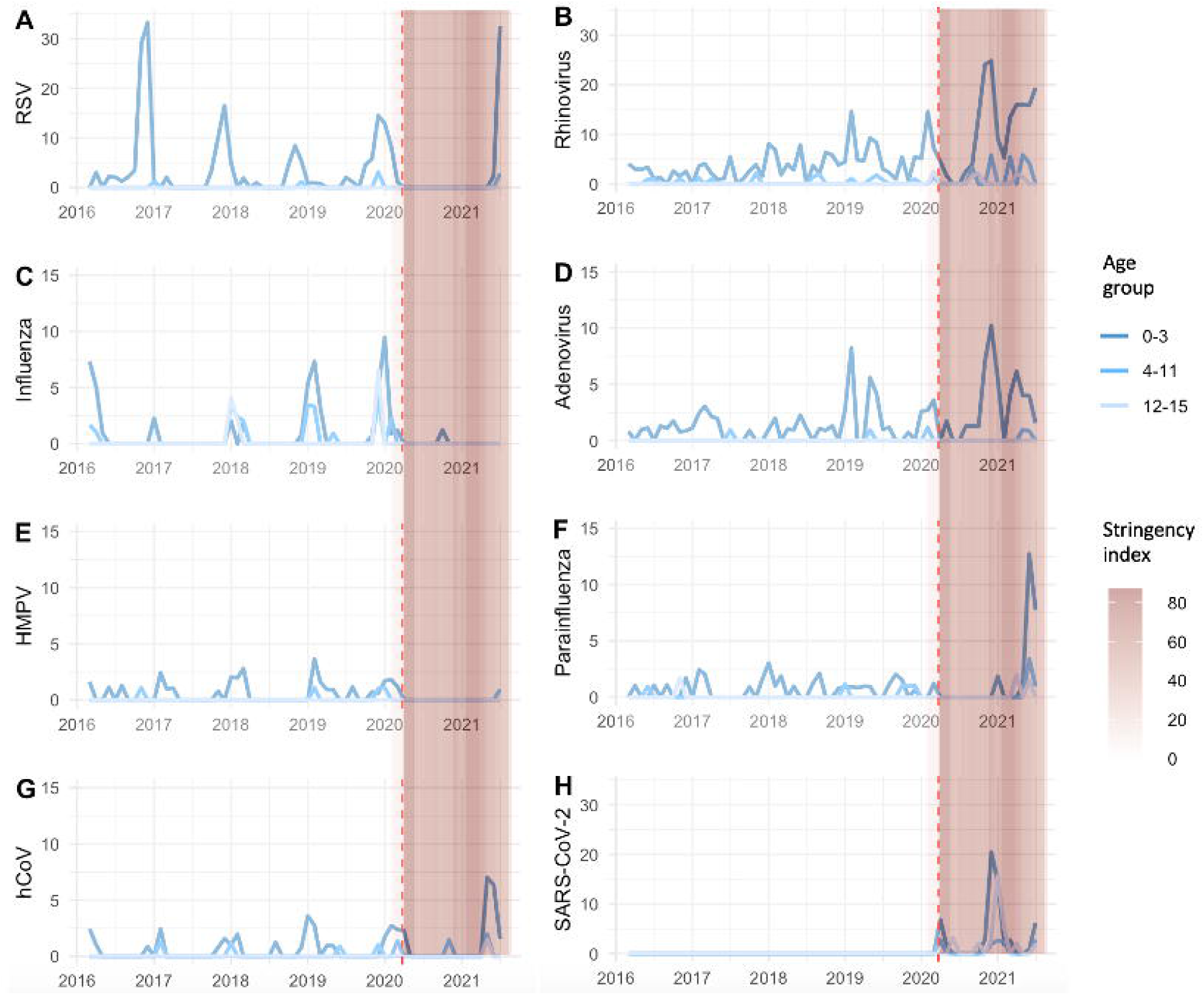
Rates of respiratory diagnoses over time, by pathogen and age group (number of positive diagnoses per 1,000 attendances per month). Vertical coloured bars represent the daily Oxford COVID-19 Government Response Tracker (OxCGRT) stringency index values on a scale from 0 to 100, with larger (darker pink) values indicating that higher stringency measures were in place in England. Red vertical dotted line indicates start of pandemic period, defined here as March 2020. RSV = respiratory syncytial virus, HMPV = human metapneumovirus, hCoV = human coronaviruses (non-SARS-CoV-2)

The patterns of suppression and resurgence during the pandemic varied by pathogen (Figure 3, Supplementary figure 2). Detection of all pathogens was suppressed during the first national lockdown and during summer 2020 (7 months). Rhinovirus (Figure 3B) and adenovirus (Figure 3D) were the first pathogens to re emerge in September 2020 with incidence rising to to 25 and 10 cases/1000 attendances/month in the 0-3 year age group respectively (a period where schools and childcare facilities were open and lockdown rules were relaxed). Rates fell in January-February 2021, during a period of increased stringency of lockdown measures, then rose again March-June 2021 to 19 and 6 cases/1000 attendances/month in the 0-3 year age group respectively, when stringency of lockdown measures was reduced. No statistically significant difference was seen in age of attendees between pandemic vs pre-pandemic periods (rhinovirus: pre-pandemic 0.7yr [IQR 0.3-1.6] vs. pandemic 1.3yr [0.7-2.1], p = 0.46 and adenovirus: pre-pandemic 1.2yr [0.6-1.6] vs. pandemic 1.3yr [1.1-1.8], p = 0.35, Chi-squared).

In contrast, RSV (Figure 3A) cases remained suppressed for the first 15 months of the pandemic, much longer than rhinovirus and adenovirus; the usual seasonal winter peak did not occur in 2020/21. In July 2021 RSV rates rose out of season in the pre-school age group (32/1,000 attendances/month in 0-3yr olds), more than double the rate seen pre-pandemic in December 2019 (14/1,000 attendances/month). The median age of RSV attendees was higher during the pandemic (pre-pandemic 0.3yr [IQR 0.2-0.9] vs. pandemic 1.8yr [IQR 0.7-2.8], p = 0.03, Chi-squared).

Influenza A/B (Figure 3C) remained suppressed throughout the pandemic period reported here, with only 1 case diagnosed. Parainfluenzavirus and hCoV rates were unusually high in May-June 2021 to 12 and 7 cases/1,000 attendances/month in the 0-3 year age group respectively (Figure 3F,G). Very few cases of metapneumovirus (Figure 3E) were seen during the pandemic, however baseline rates are usually low.

### Co-infections

Of the 935 ED attendances where a positive respiratory virus test was obtained, 770 were mono-infected and 165 co-infected (134 with two pathogens, 27 with three pathogens and 4 with four pathogens). Co-infections were eliminated by the first national lockdown. Co-infections took longer (2 months) to re-emerge after the relaxation of restrictions than mono-infections. No four-pathogen co-infections occurred during the pandemic (Figure 4).

**Figure 4:**
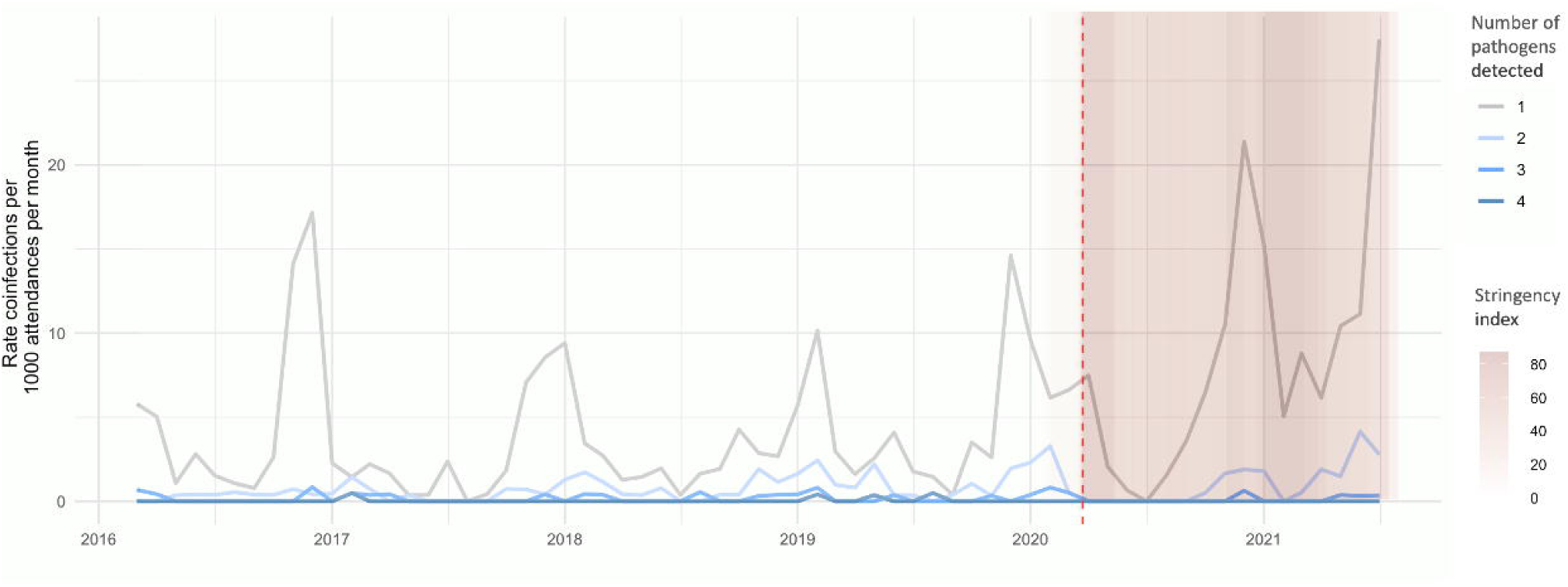
Rate of respiratory virus co-infections over time in paediatric ED attenders. Vertical coloured bars represent the daily Oxford COVID-19 Government Response Tracker (OxCGRT) stringency index values on a scale from 0 to 100, with larger (darker pink) values indicating that higher stringency measures were in place in England. Red vertical dotted line indicates start of pandemic period, defined here as March 2020.

### Disease severity

Partial PEWS were calculated for 571 attendances with positive respiratory virus PCRs. There were no significant differences in PEWS between attendees diagnosed with different respiratory viral pathogens pre-pandemic (p=0.27, Kruskal-Wallis). There was no significant difference between PEWS pre- and during the pandemic (p=0.24, Mann-Witney U test). A higher PEWS was seen for attendees with adenovirus during the pandemic compared to pre-pandemic (p=0.04, Mann-Witney U test), no other differences in PEWS between pre-pandemic v.s. pandemic periods were seen for other pathogens (all p-values >0.1, Mann-Witney U test) (Figure 5). There were fewer deaths within 14 days of a positive respiratory virus PCR during the pandemic than before the pandemic (0.3 vs. 0.8 deaths per 100 positive PCRs respectively, p <0.0001).

**Figure 5:**
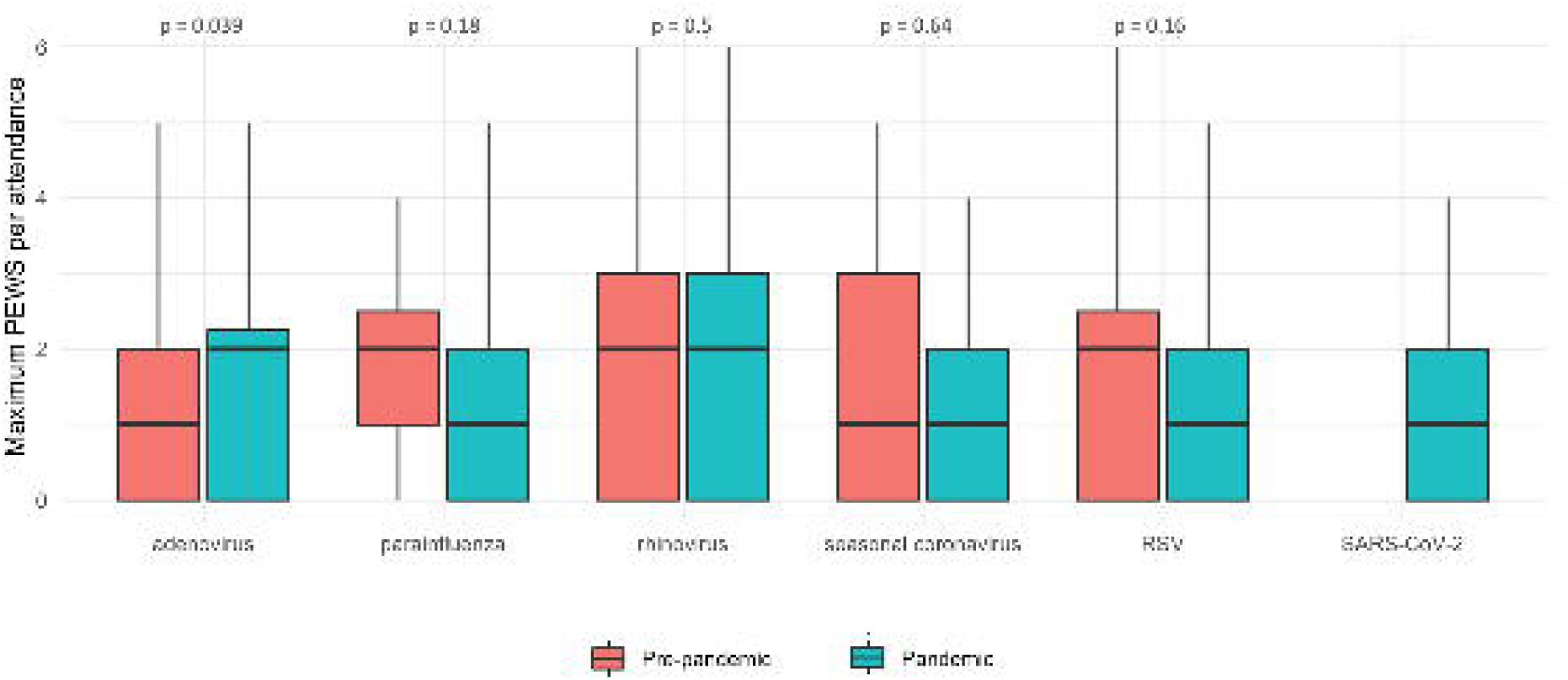
Paediatric early warning scores (PEWS) comparison between pathogens and time periods. Maximum PEWS per ED attendance comparing pre-pandemic and pandemic periods for the five respiratory viruses detected during both time periods of the study, plus SARS-CoV-2 for reference. The central bar indicates the median PEWS, the lower and upper bounds of the box indicate the first and third quartiles (IQR), the lower whisker extends from the first quartile to the lowest value within 1.5^*^IQR of the first quartile, the upper whisker extends from the third quartile to the highest value within 1.5^*^IQR of the third quartile. P values (Mann-Witney U test) comparing pre-pandemic and pandemic PEWS for each pathogen are presented above the paired bars.

## DISCUSSION

Major changes in the incidence of paediatric viral RTI occurred in Oxfordshire during the pandemic, similarly to the rest of the UK. Following an initial period of low incidence for all pathogens during the first national lockdown, pathogen-dependent patterns of resurgence were seen from September 2020 onwards, with early resurgence of rhinovirus and adenovirus, and a delayed inter-seasonal resurgence of RSV. Although RSV detection rates in July 2021 were high, and occurred in older infants, cases were not clinically more severe.

Oxfordshire data pre-pandemic is representative of seasonal paediatric viral infections, with seasonal winter fluctuations of RSV and influenza(26). Seasonal winter patterns were eliminated during the first UK lockdown period and incidence remained low, as had been the case across Europe (with the exception of France and Iceland)(27), until an inter-seasonal spike in RSV incidence in Oxfordshire in July 2021.

Similar inter-seasonal RSV resurgences have been seen in the UK, Europe and countries in the Southern Hemisphere(9,15,21–23), also in older children(27,29–31) (likely due to lack of pre-existing immunity in 1-2 year olds due to decreased exposure), raising challenges for management of varying presentations of RSV in this older age group. It has been hypothesised that nursery and primary school closures have an impact on RSV transmission due to predominance in children under 5 years old; France and Iceland both had policies of keeping primary schools and daycare facilities open(27). In Oxfordshire, the inter-seasonal RSV spike occurred just as schools closed for the summer holidays, alongside presentations occurring predominantly in pre-school aged children, this raises the question whether daycare settings for younger children (which largely remain open over the summer) are the primary source of transmission, with case numbers fueled by easing of lockdown measures within the general population, parents returning to work and easing of COVID restrictions in daycare settings.

This inter-seasonal RSV peak required a change of testing strategy with an introduction of out-of-season RSV testing and an extension of monthly preventative palivizumab for infants at risk for severe RSV disease(26,28), and contributed to an uncharacteristic summer peak in paediatric ED attendances and pressure on staffing. Influenza remained suppressed throughout, a pattern seen globally(32), presenting challenges for the selection of vaccine strains for the future winter influenza vaccination campaign.

In contrast, less seasonal variability was seen with rhinoviruses pre-pandemic; although their relative prevalence decreases in winter due to influenza interference, they are usually the most prevalent respiratory viral agent during summer months (33,34). Rhinovirus incidence increased to rates above those seen pre-pandemic in September-December 2020 (a period where schools and daycare were open and lockdown rules were relaxed), fell in January-February 2021, and rose again March-May 2021.

Similar rhinovirus resurgences were seen in Australia(35), Germany(18), New Zealand(19), Japan(20) and in adults in England(17). A rapid rise in adenovirus, similar to that seen for rhinoviruses after school re-opening, was seen in this study. HCoV show a winter seasonal pattern pre-pandemic, with a rise in detection in May-June 2021. Parainfluenza virus patterns were inconsistent pre-pandemic, rising in June 2021. SARS-CoV-2 incidence reflects the national pandemic infection curves. Reassuringly, no increase in severity (measured by PEWS) was seen in the pandemic period, except for adenovirus.

Although it is clear that social distancing measures and school/daycare closures dramatically decreased the incidence of all respiratory viruses at the start of the pandemic, it is difficult to disaggregate the effect of social distancing in general vs. the specific impact of school closures thereafter. It is interesting to note that whilst rhinovirus and adenovirus cases tend to fluctuate with school openings, the rise in RSV cases in July 2021 occurred at the point when schools were closing for the summer holidays. Furthermore, the majority of cases in this study are in pre-school aged children, likely a combination of a true high incidence in this age group (as is usually the case) and a higher likelihood of severe illness in pre-school age children requiring hospital attendance. Therefore paradoxically, the greatest benefit of school closures and lockdown might be reducing community incidence of respiratory viruses and therefore acquisition and hospitalisation in the pre-school age group. An important piece of future work will be to understand which components of the public health interventions were most effective for preventing various respiratory virus infection and hospitalisation in different age groups, in particular understanding the contribution of school closures, given the many negative effects of closing schools on children and on society.

There are several hypotheses exploring the different patterns of resurgence between viral pathogens, including differences in the durability of immunity and the impact of reduced exposures on natural “boosting”, respiratory virus “interference” in which one epidemic delays the start or accelerates the end of the other viral epidemic (as was seen in the 2009 influenza pandemic in which the RSV epidemic was also delayed)(36–38), and differences in viral structure (for example the presence of an envelope) or transmission altering susceptibility to social distancing and inactivation by handwashing and surface cleaning.

There were limitations to this study. Respiratory diagnoses amongst ED attendees represent the more severe end of the disease spectrum and although these reflect community prevalence (this study shows similar trends to Public Health England surveillance data(39)), they likely over-represent diagnoses in the pre-school children more likely to require hospital care for respiratory infection. Furthermore, some children, for example those with croup who are not conventionally sampled, are not represented in the dataset. Changes in healthcare seeking behaviours during the pandemic impact the calculated rates of infection, however this is partly mitigated by presenting respiratory diagnoses per 1000 attendances. The increased incidence of rhinovirus, adenovirus, hCoV and parainfluenza in the pandemic relative to pre-pandemic in this study is likely exaggerated by an ascertainment bias, due to increased use of the more comprehensive Biofire respiratory pathogen panel test for deteriorating patients or those requiring aerosol generating procedures during the pandemic and introduction of quadruple Influenza A/B/RSV/SARS-CoV-2 admission screening in July 2021.

Although we have divided the study period into “pandemic” and “pre-pandemic” periods, SARS-CoV-2 circulated in the UK during the early weeks of 2020, defined here as “pre-pandemic”. Since not all periods of lockdown are equivalent in terms of stringency, we used the Oxford COVID-19 government response tracker’s Stringency Index(24) to indicate the degree of restrictions in place throughout the study.

However this stringency index does not capture population adherence to lockdown and social distancing measures.

Further studies, including the ‘Bronchstart’ study(40), will be required to assess the ongoing impact of the COVID-19 pandemic on respiratory virus incidence in both paediatric and adult populations, to inform workforce planning to adapt to changing seasonal patterns of service demand, altered testing strategies and use of anti-viral prophylaxis in high risk groups. We also need to understand which public health interventions are most effective for different pathogens, and assess whether any ongoing social distancing measures (for example public health messaging about hand washing, social distancing, ventilation and masking) could be useful to reduce the burden of RTI in the long term.

## Data Availability

The epidemiological datasets analysed are not publicly available as they contain personal data but are available from the Infections in Oxfordshire Research Database (https://oxfordbrc.nihr.ac.uk/research-themes-overview/antimicrobial-resistance-and-modernising-microbiology/infections-in-oxfordshire-research-database-iord/), subject to an application and research proposal meeting the ethical and governance requirements of the Database.

## ACKNOWLEDGEMENTS

We thank all the people of Oxfordshire who contribute to the Infections in Oxfordshire Research Database. Research Database Team: L Butcher, H Boseley, C Crichton, DW Crook, DW Eyre, O Freeman, J Gearing (community), R Harrington, K Jeffery, M Landray, A Pal, TEA Peto, TP Quan, J Robinson (community), J Sellors, B Shine, AS Walker, D Waller. Patient and Public Panel: G Blower, C Mancey, P McLoughlin, B Nichols

