## Supplementary materials for "Changes in paediatric respiratory infections at a UK teaching hospital 2016-2021; impact of the SARS-CoV-2 pandemic"

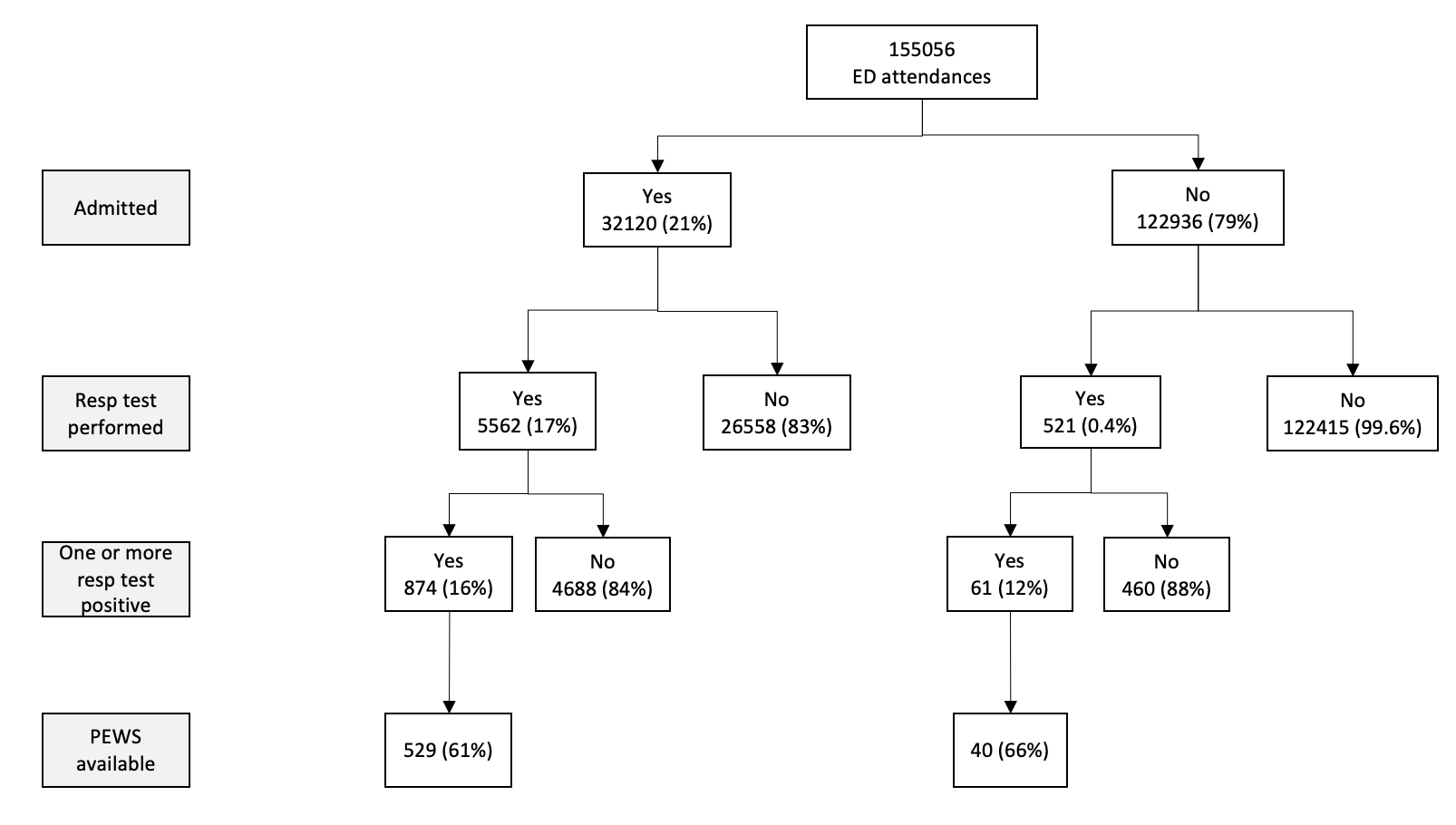

**Supplementary figure 1: Flowchart showing the number of attendances, admissions, testing and positivity rates, and vital signs for the study cohort**

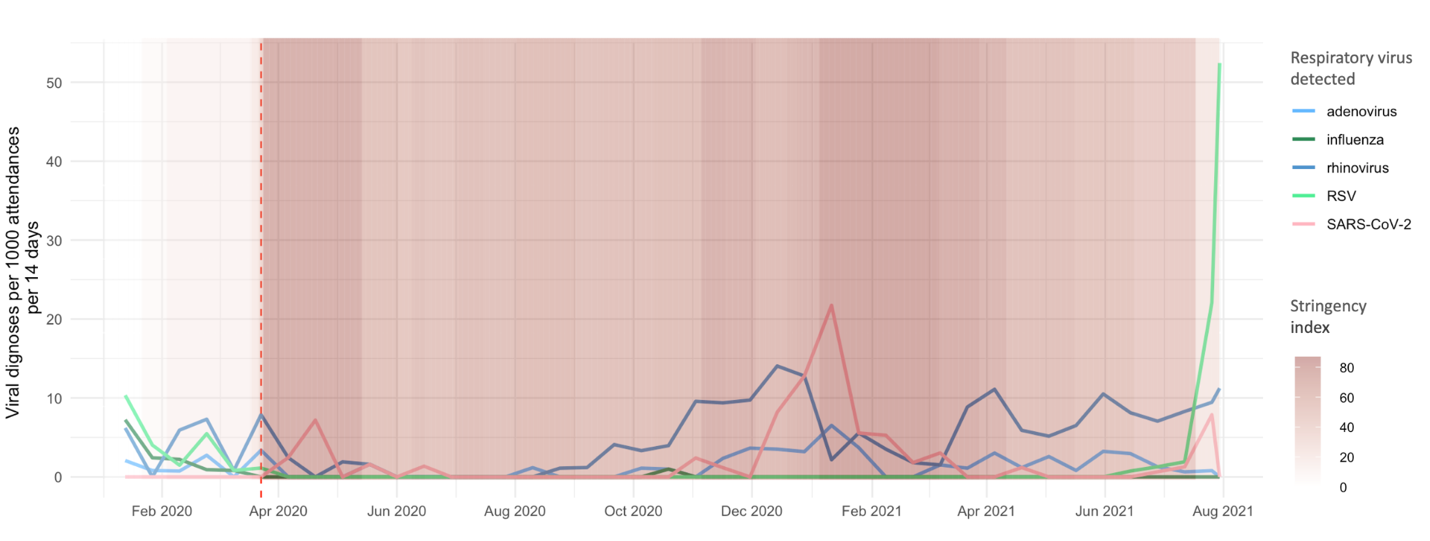

**Supplementary figure 2:  Rates of respiratory diagnoses over time, by pathogen during 2020/21 (number of positive diagnoses per 1,000 attendances per 14 days).** Highlighting differences in resurgence of 5 key respiratory viruses in relation to changes in social distancing stringency. Red vertical dotted line indicates the start of the pandemic period. Vertical coloured bars represent the daily Oxford COVID-19 Government Response Tracker (OxCGRT) stringency index values on a scale from 0 to 100, with larger (darker pink) values indicating that higher stringency measures were in place in England.

| **Test** | **PCR platform(s) used** |
| --- | --- |
| Influenza A/B/RSV PCR | Cepheid Xpert® Xpress Flu/RSV |
| Biofire respiratory PCR | Biofire FilmArray multiplex respiratory PCR (RP2 and RP2.1) |
| SARS-CoV-2 PCR | Public Health England SARS-CoV-2 assay  Abbott RealTime SARS-CoV-2  Cepheid Xpert® Xpress SARS-CoV-2  Cepheid Xpert® Xpress SARS-CoV-2/Flu/RSV  ABI 7500 platform with with the US Centers for Disease Control and Prevention Diagnostic Panel  Thermo Fisher TaqPath assay |
| Influenza A/B/RSV and SARS-CoV-2 PCR | Cepheid Xpert® Xpress SARS-CoV-2/Flu/RSV |

**Supplementary table 1: PCR platforms used for detection of respiratory viruses.**

|  | **Overall**  **(n= 155056)** | **Respiratory virus test performed** | | **P-value (comparing tested vs. untested)** |
| --- | --- | --- | --- | --- |
|  |  | **Yes**  **(n=6083)** | **No**  **(n=148973)** |  |
| **Age group in years**    0-3; pre-school    4-11; primary school    12-15; secondary school | 64096 (41.3%)  58734 (37.9%)  32226 (20.7%) | 3453 (56.8%)  1687 (27.7%)  943 (15.5%) | 60643 (40.7%)  57047 (38.3%)  31283 (30.0%) | <0.0001 |
| **Sex**    Male    Female    Not stated | 87010 (56.1%)  68044 (43.9%)  2 (<0.1%) | 3426 (56.3%)  2657 (43.7%)  0 | 83584 (56.1%)  65387 (43.9%)  2 (<0.1%) | 0.98 |
| **Ethnicity**   White   Asian   Mixed   Black   Chinese   Other   Not Stated | 118314 (76.3%)  10005 (6.5%)  7537 (4.9%)  3250 (2.1%)  1029 (0.7%)  2876 (1.9%)  12045 (7.8%) | 4552 (74.8%)  470 (7.7%)  309 (5.1%)  142 (2.3%)  37 (0.6%)  131 (2.2%)  442 (7.3%) | 113762 (76.4%)  9535 (6.4%)  7228 (4.9%)  3108 (2.1%)  992 (0.7%)  2745 (1.8%)  11603 (7.8%) | 0.0003 |
| **IMD Score** | 11.7 (7.1 - 20.1) | 11.3 (6.7 - 19.7) | 11.7 (7.1 - 20.4) | <0.0001 |

**Supplementary table 2: Demographics of ED attendees with/without respiratory virus testing.** Median (IQR) or frequency (%) provided. P-values calculated with Pearson’s Chi-squared test for categorical data and Mann-Whitney U test for numeric data. IMD = index of multiple deprivation; PCR = polymerase chain reaction

|  | **Tests performed** | | | |
| --- | --- | --- | --- | --- |
|  | **Flu A/B/RSV** | **Biofire** | **Biofire +**  **Flu A/B/RSV** | **Biofire +**  **SARS-CoV-2** |
| Total | 290 | 588 | 1 | 3 |
| Number of monoinfections | 288 | 426 | 0 | 0 |
| Number of viral coinfections | 2 | 162 | 1 | 3 |
| Breakdown by respiratory virus | | | | |
| MONOINFECTIONS | | | | |
| RSV | 219 | 33 | 0 | 0 |
| Influenza A | 51 | 9 | 0 | 0 |
| Influenza B | 18 | 3 | 0 | 0 |
| Human rhinovirus/enterovirus | NA | 265 | 0 | 0 |
| Adenovirus | NA | 38 | 0 | 0 |
| Metapneumovirus | NA | 19 | 0 | 0 |
| Parainfluenza 1 | NA | 2 | 0 | 0 |
| Parainfluenza 2 | NA | 2 | 0 | 0 |
| Parainfluenza 3 | NA | 29 | 0 | 0 |
| Parainfluenza 4 | NA | 4 | 0 | 0 |
| HCoV NL63 | NA | 12 | 0 | 0 |
| HCoV OC43 | NA | 5 | 0 | 0 |
| HCoV HKU1 | NA | 2 | 0 | 0 |
| HCoV 229E | NA | 1 | 0 | 0 |
| Parechovirus | NA | 1 | 0 | 0 |
| Bocavirus | NA | 1 | 0 | 0 |
| DUAL INFECTIONS | | | | |
| Rhinovirus + adenovirus | NA | 44 | 0 | 0 |
| Rhinovirus + RSV | NA | 18 | 0 | 0 |
| Rhinovirus + parainfluenza 3 | NA | 12 | 0 | 0 |
| Rhinovirus + metapneumovirus | NA | 8 | 0 | 0 |
| Rhinovirus + HCoV HKU | NA | 6 | 0 | 0 |
| Adenovirus + 229E | NA | 3 | 0 | 0 |
| Adenovirus + parainfluenza 3 | NA | 3 | 0 | 0 |
| Parainfluenza 2 + 3 | NA | 3 | 0 | 0 |
| Parainfluenza 3 + HCoV NL63 | NA | 3 | 0 | 0 |
| Rhinovirus + parainfluenza 4 | NA | 3 | 0 | 0 |
| Rhinovirus + parainfluenza 1 | NA | 3 | 0 | 0 |
| Adenovirus + metapneumovirus | NA | 2 | 0 | 0 |
| Rhinovirus + bocavirus | NA | 2 | 0 | 0 |
| Rhinovirus + HCoV NL63 | NA | 2 | 0 | 0 |
| Rhinovirus + Influenza A | NA | 2 | 0 | 0 |
| Adenovirus + bocavirus | NA | 1 | 0 | 0 |
| Adenovirus + HCoV HKU | NA | 1 | 0 | 0 |
| Adenovirus + HCoV OC43 | NA | 1 | 0 | 0 |
| Adenovirus + Influenza A | NA | 1 | 0 | 0 |
| Adenovirus + parainfluenza 4 | NA | 1 | 0 | 0 |
| Adenovirus + RSV | NA | 1 | 0 | 0 |
| HCoV 229E + bocavirus | NA | 1 | 0 | 0 |
| HCoV OC43 + parainfluenza 3 | NA | 1 | 0 | 0 |
| Influenza A + B | 0 | 1 | 0 | 0 |
| Influenza A + parainfluenza 2 | NA | 1 | 0 | 0 |
| Influenza A + RSV | 2 | 1 | 0 | 0 |
| Influenza B + bocavirus | NA | 1 | 0 | 0 |
| Metapneumovirus + bocavirus | NA | 1 | 0 | 0 |
| Parainfluenza 1 + metapneumovirus | NA | 1 | 0 | 0 |
| Parainfluenza 1+4 | NA | 1 | 0 | 0 |
| Rhinovirus + HCoV 229E | NA | 1 | 0 | 0 |
| Rhinovirus + HCoV OC43 | NA | 1 | 0 | 0 |
| Rhinovirus + parechovirus | NA | 1 | 0 | 0 |
| RSV + HCoV HKU | NA | 1 | 0 | 0 |
| RSV + parainfleunza 3 | NA | 1 | 0 | 0 |
| RSV + parainfluenza 2 | NA | 1 | 0 | 0 |
| Rhinovirus + SARS-CoV-2 | NA | 0 | 0 | 2 |
| TRIPLE INFECTIONS | | | | |
| Rhinovirus + adenovirus + HCoV NL63 | NA | 2 | 0 | 0 |
| Rhinovirus + adenovirus + HCoV OC43 | NA | 2 | 0 | 0 |
| Rhinovirus + adenovirus + RSV | NA | 2 | 0 | 0 |
| Rhinovirus + RSV + HCoV HKU | NA | 2 | 0 | 0 |
| Rhinovirus + adenovirus + bocavirus | NA | 1 | 0 | 0 |
| Rhinovirus + adenovirus + HCoV HKU | NA | 1 | 0 | 0 |
| Rhinovirus + adenovirus + influenza A | NA | 1 | 0 | 0 |
| Rhinovirus + adenovirus + metapneumovirus | NA | 1 | 0 | 0 |
| Rhinovirus + bocavirus + HCoV NL63 | NA | 1 | 0 | 0 |
| Rhinovirus + boca virus+ HCoV OC43 | NA | 1 | 0 | 0 |
| Rhinovirus + metapneumovirus + bocavirus | NA | 1 | 0 | 0 |
| Rhinovirus + metapneumovirus + HCoV NL63 | NA | 1 | 0 | 0 |
| Rhinovirus + metapneumovirus + HCoV OC43 | NA | 1 | 0 | 0 |
| Rhinovirus + parainfluenza 3 + boca | NA | 1 | 0 | 0 |
| Rhinovirus + parainfluenza 3 + HCoV NL63 | NA | 1 | 0 | 0 |
| Rhinovirus + parainfluenza 4 + influenza A | NA | 1 | 0 | 0 |
| Rhinovirus + parechovirus + bocavirus | NA | 1 | 0 | 0 |
| Rhinovirus + parechovirus + HCoV NL63 | NA | 1 | 0 | 0 |
| Rhinovirus + RSV + HCoV OC43 | NA | 1 | 0 | 0 |
| Rhinovirus + RSV + HCoV 229E | NA | 1 | 0 | 0 |
| Rhinovirus + RSV + metapneumovirus | NA | 1 | 0 | 0 |
| Adenovirus + parainfluenza 3 + HCoV 229E | NA | 1 | 0 | 0 |
| RSV + HCoV HKU + HCoV OC43 | NA | 0 | 1 | 0 |
| Rhinovirus + adenovirus + SARS-CoV-2 | NA | 0 | 0 | 1 |
| QUADRUPLE INFECTIONS | | | | |
| Rhinovirus + adenovirus + metapneumovirus + parainfluenza 3 | NA | 1 | 0 | 0 |

**Supplementary table 3: Breakdown of pathogens detected by test type, in cases where more than one respiratory virus was tested for.**
